# Acceptability and feasibility of digital adherence technologies for tuberculosis treatment supervision: A meta-analysis of implementation feedback

**DOI:** 10.1101/2023.01.26.23284950

**Authors:** Kevin Guzman, Rebecca Crowder, Anna Leddy, Noriah Maraba, Lauren Jennings, Shahriar Ahmed, Sonia Sultana, Baraka Onjare, Lucas Shilugu, Jason Alacapa, Jens Levy, Achilles Katamba, Alex Kityamuwesi, Aleksey Bogdanov, Kateryna Gamazina, Adithya Cattamanchi, Amera Khan

**Author notes:** **Corresponding Authors: Amera, Khan, DrPH^8^**, Global Health Campus, Chemin du Pommier 40, 1218 Geneva, Switzerland, **Adithya Cattamanchi, MD^1^**, 1001 Potrero Ave, San Francisco, CA, 94110 USA. senior co-authors (Adithya Cattamanchi, Amera Khan).

## Abstract

**Introduction:** Digital adherence technologies (DATs) have emerged as an alternative to directly observed therapy (DOT) for supervisions of tuberculosis (TB) treatment. We conducted a meta-analysis of implementation feedback obtained from people with TB and health care workers (HCWs) involved in TB REACH Wave 6-funded DAT evaluation projects.

**Methods:** Projects administered standardized post-implementation surveys based on the Capability, Opportunity, Motivation, Behavior (COM-B) model to people with TB and their health care workers. The surveys included questions on demographics and technology use, Likert scale questions to assess capability, opportunity, and motivation to use DAT and open-ended feedback. We summarized demographic and technology use data descriptively, generated pooled estimates of responses to Likert scale questions within each COM-B category for people with TB and health care workers using random effects models, and performed qualitative analysis of open-ended feedback using a modified framework analysis approach.

**Results:** The analysis included surveys administered to 1290 people with TB and 90 HCWs across 6 TB REACH-funded projects. People with TB and HCWs had an overall positive impression of DATs with pooled estimates between 4·0 to 4·8 out of 5 across COM-B categories. However, 44% of people with TB reported taking TB medications without reporting dosing via DATs and 23% reported missing a dose of medication. Common reasons included problems with electricity, network coverage, and technical issues with the DAT platform. DATs were overall perceived to reduce visits to clinics, decrease cost, increase social support, and decrease workload of HCWs.

**Conclusion:** DATs were acceptable in a wide variety of settings. However, there were challenges related to the feasibility of using current DAT platforms. Implementation efforts should concentrate on ensuring access, anticipating, and addressing technical challenges, and minimizing additional cost to people with TB.

**Author Summary:** Digital adherence technologies (DATs) are increasingly being implemented as an alternative to traditional directly observed therapy (DOT) for TB treatment. However, to date there are limited data on their feasibility and acceptability among both persons on treatment and health care workers, resulting in only a conditional recommendation for their use in TB treatment by the World Health Organization in their 2017 *Guidelines for treatment of drug-susceptible tuberculosis and patient care*.

Our study provides information on the feasibility and acceptability of implementing and using different DATs in a variety of settings and target populations for TB treatment adherence. The use of a similar survey across multiple sites helps provides a common understanding of facilitators and barriers on the use of DATs as global and national TB programs consider the expansion of the use of these tools. Our evidence demonstrates a high acceptability of DATs and supports further implementation of DATs as a component of TB treatment support. However, implementation efforts need to address issues concerning access to these tools, the technical challenges that are associated with the platforms, while minimizing additional burdens and costs to people with TB.

## Introduction

Despite the availability of effective treatment, tuberculosis (TB) remains a leading cause of death from an infectious agent, behind only COVID-19.^1^ Successful treatment of drug-susceptible TB (DS-TB) requires high levels of adherence to treatment lasting at least 4 months. Since the 1990s, directly observed therapy (DOT) has been employed globally to monitor and support adherence to TB medications. However, there is limited evidence that DOT improves treatment completion in comparison to self-administered therapy, ^2.3^ and treatment completion rates remain below the 90% target in most high burden countries. ^1^ DOT is associated with catastrophic costs and rigid enforcement of DOT may conflict with the autonomy, dignity, and integrity of people with TB. ^4,5^

There has been increasing interest in digital adherence technologies (DATs) as a more person-centered alternative to DOT. DATs enable people with TB to take TB medicines at a time and place of their choosing and allow health care workers (HCWs) to monitor adherence and provide tailored adherence support to those who need it. ^6^ Several different commercial DAT platforms have been deployed in high TB burden countries. These include 99DOTS, a phone-based technology by which people self-report adherence via calls or texts to phone numbers that are revealed behind pills in a medication sleeve; ^7^ electronic smart pillboxes made of either plastic or cardboard, such as evriMED, which passively register dosing events when the pillbox is opened; ^8^ and video directly observed therapy (VDOT), whereby people record and upload videos documenting medication ingestion using a smartphone or computer with a camera.^9^ Although these technologies are increasingly being implemented, there are limited data on their effectiveness, acceptability, and feasibility, resulting in only a conditional recommendation for their use in TB treatment by the WHO.^10^

To assess the acceptability and feasibility of DATs from a variety of settings and target populations, the Stop TB Partnership’s TB REACH initiative funded 14 DAT projects in 12 countries as part of their Wave 6 funding portfolio. ^11^ We analyzed standardized implementation feedback obtained from people with TB and HCWs as part of the TB REACH-funded projects.

## Methods

### Studies and participants

We excluded TB REACH Wave 6-funded DAT evaluation projects that did not use a commercial DAT platform, share individual participant data with the Stop TB Partnership or exclusively enrolled people with drug-resistant TB (DR-TB). In addition, we excluded people with TB from the analysis if they were younger than 15 years old or were missing data on sex and excluded HCWs from the analysis if they did not provide direct care to people on TB treatment.

### Survey design

Standardized post-implementation surveys for people with TB and HCWs were designed by the TB REACH DAT working group to assess the feasibility and acceptability of using a DAT as part of TB treatment. Feasibility was defined as the extent to which DAT could be successfully used or carried out within a given setting.^12^ Acceptability was defined as the perception that the DAT was agreeable, palatable, or satisfactory.^13^ To assess feasibility and acceptability, the survey included questions about access to technology and experience using DATs and Likert scale questions, developed to reflect selected domains of the Theoretical Domains Framework (TDF).^14^ The TDF integrates 33 psychologic theories relevant to behavior change into 14 domains related to Capability (knowledge, attention, memory, and decision processes necessary to use DAT), Opportunity (social and environmental context conducive to using DAT), or Motivation (optimism, reinforcement, and emotion to want to use DAT) to perform a Behavior (COM-B).^15,16^ Health worker surveys also included constructs from the Unified Theory of Acceptance and Use of Technology (UTAUT), a consolidated framework to explain information systems usage behavior.^17–19^ In addition, the surveys included open-ended questions to further explore DAT feasibility and acceptability, as well as recommendations for improvement (see *Supplemental Tables S1– S2)*.

Projects administered surveys to a random sample of people with TB and HCWs. In some cases, projects modified the survey tools to exclude certain questions or include additional questions. Survey administration varied by project and surveys were conducted either by HCWs or research staff. Questionnaires were translated into the local language as needed. This analysis included only survey questions that were used in common by all projects.

### Data analysis

Responses from people with TB and HCWs were analyzed separately and stratified by DAT platform evaluated. Demographics and data on access to and use of technology were summarized descriptively. Responses to Likert scale questions were summarized by project using means and standard deviations. A response of agree or strongly agree reflected a favorable impression of DAT. If the question was asked such that agreement indicated an unfavorable impression of DAT, responses were reversed prior to analysis. Composite scores for capability, opportunity, and motivation were created for each person with TB and HCW by calculating the mean response to Likert scale questions within that category.

Pooled estimates for capability, opportunity, and motivation scores across projects were generated using random effects, sample size weighted models. A sample size weighted model was chosen over an inverse variance weighted model due to lack of variance in survey responses across participants for some projects. Secondary analyses assessed differences by gender and age category (≥55 years old vs. <55 years old). Differences in mean capability, opportunity, and motivation scores were calculated by gender and by age category for each project. Pooled estimates of the gender and age category differences in mean scores were then generated using the random effects, sample size weighted model described above. Stata 15 was used for all quantitative analyses.

Qualitative analysis of responses to open ended survey questions was performed using a modified framework analysis approach.^20^ We began by sorting responses into frames reflecting the open-ended questions posed in each survey. Content that was common across participants (mentioned by four or more participants) or that was uncommon but offered unique insights was flagged for inclusion as a theme. Themes that emerged across frames were considered cross-cutting themes. The main themes across the data sources were then consolidated and synthesized into chart format and organized as barriers/facilitators to DAT uptake by COM-B category, DAT type, project country and sex. This enabled us to identify similarities and differences in barriers and facilitators to DAT uptake across DAT type, country, and sex.

## Results

### Setting and study population

Of the 14 TB REACH Wave 6-funded DAT evaluation projects, 8 were excluded (2 exclusively enrolled people with drug-resistant TB, 2 used a locally-developed rather than commercial DAT platform, and 4 did not share individual data from persons on TB treatment or HCWs). The remaining six projects administered surveys to 1,316 people with TB and 92 HCWs using 99DOTS or evriMED for TB treatment supervision. Survey data from 1290 (98·0%) people with TB and 90 (97·8%) HCWs were analyzed (*Figure 1*).

**Figure 1:**
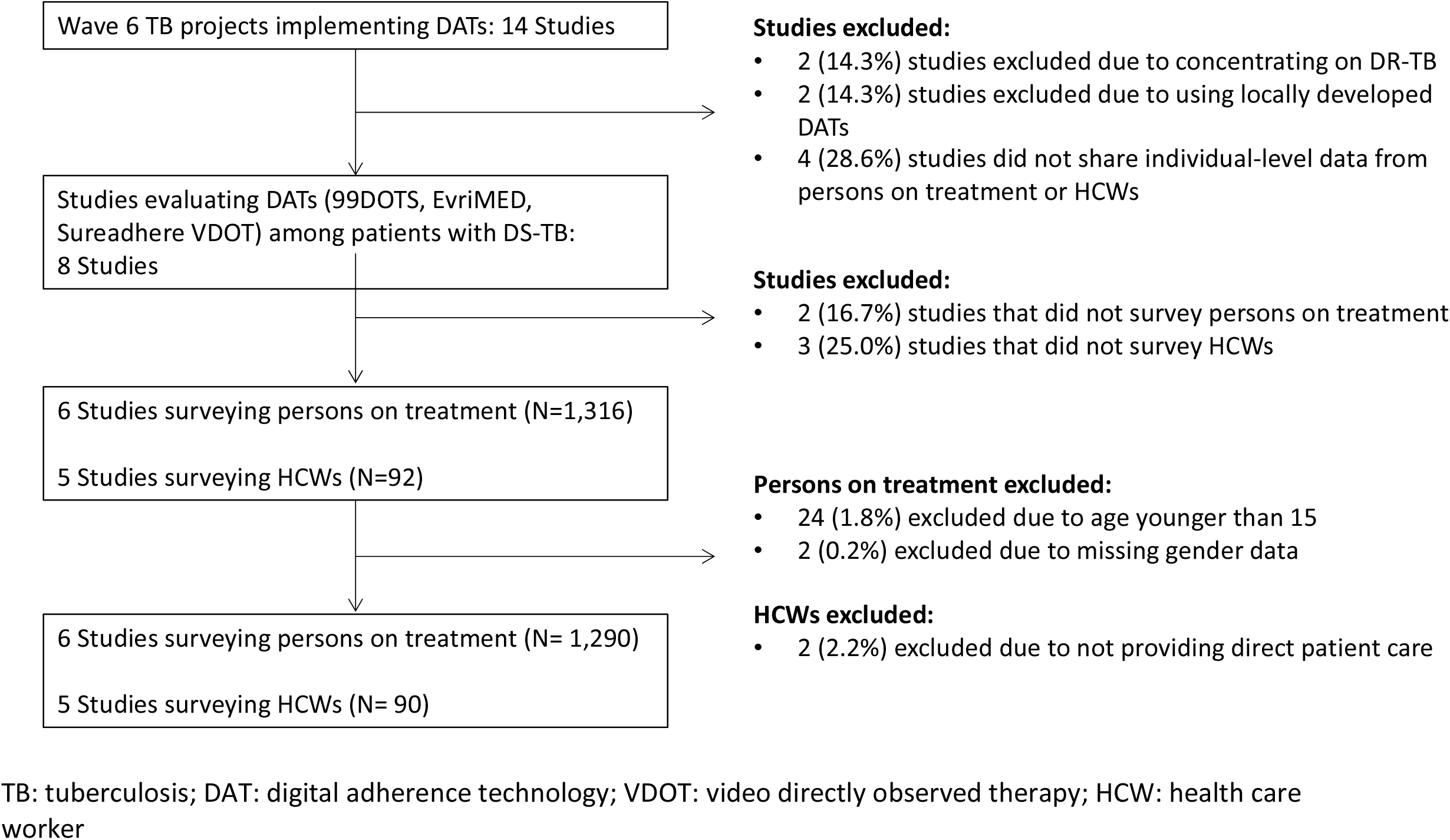
Studies and population included in meta-analysis.

Details of study setting and DAT implementation for the six included projects are provided in *Supplemental Table S3*. Projects in Uganda, South Africa, Tanzania, and Ukraine were conducted at government-run TB treatment facilities, whereas projects in Bangladesh and the Philippines implemented DATs in private sector settings. Most HCWs treated only people with drug susceptible TB, but HCWs surveyed in Ukraine were treating both people with drug susceptible and drug resistant TB.

Projects implementing 99DOTS provided medication blister packs placed within an envelope that revealed toll-free numbers when medications were pushed out of the blister pack. People with TB were instructed to call or text the toll-free numbers daily to confirm dosing. In Tanzania, however, they were charged for calls made to 99DOTS and excluded from the project if they did not have a daily minimum phone balance prior to joining the study. Projects implementing evriMED provided a smart pill box that contained TB medication blister packs. In all projects, HCWs could access and monitor daily adherence data using a mobile phone or web-based application.

In total, 1,017 people with TB using 99DOTS in 4 countries and 273 using evriMED in 2 countries completed surveys from September 2019 to May 2020 (*Table 1)*. Almost half were from Bangladesh (n=599, 46·4%). Sample sizes for the other five projects ranged from 106 to 200 people with TB. Responses to open-ended survey questions were provided by 306 people with TB in Tanzania and the Philippines using 99DOTS, and 19 in Ukraine using evriMED (*Supplemental Tables S4 – S6*).

**Table 1:**
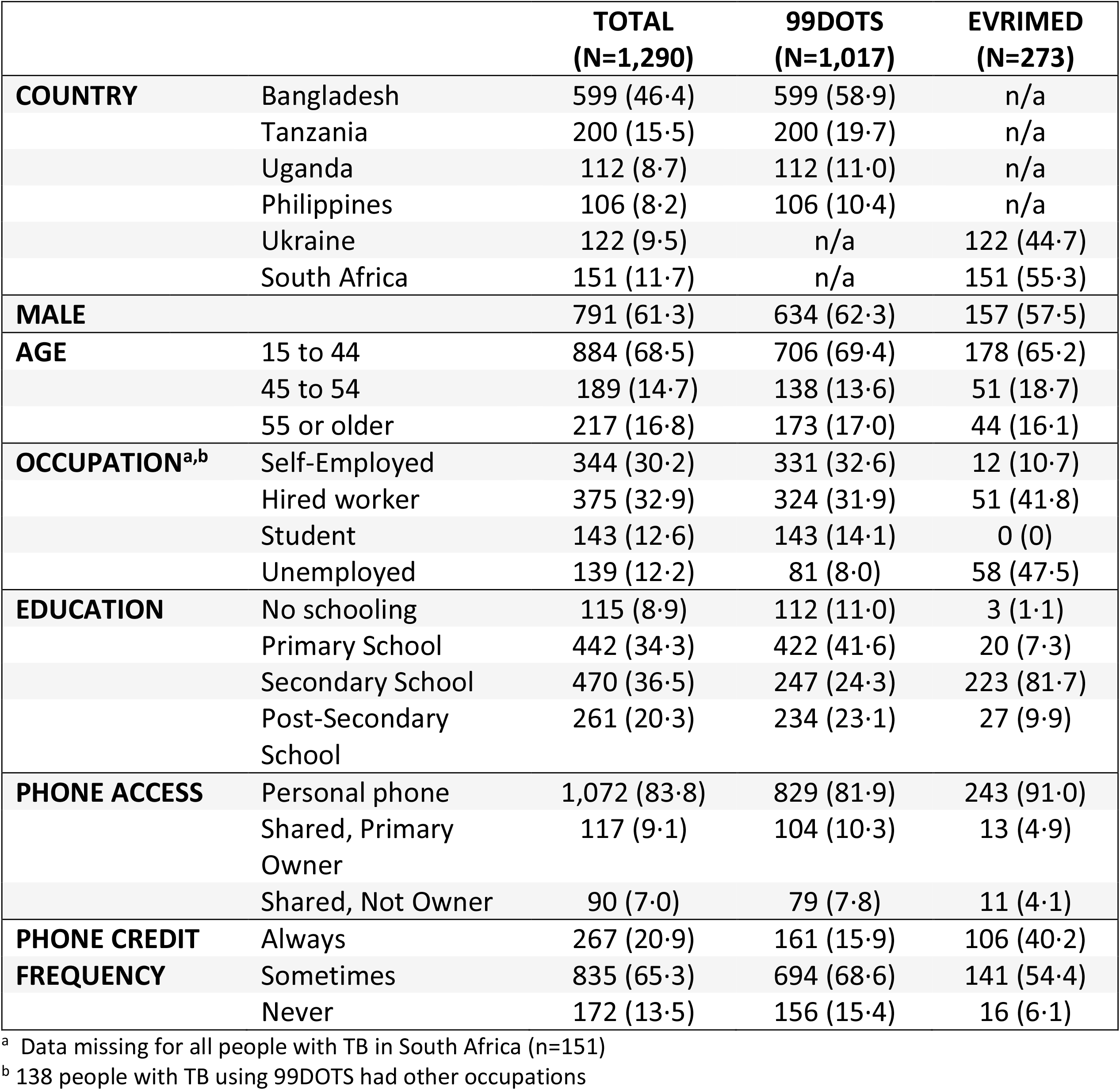
Demographic characteristics and access to technologies among persons on TB treatment.

Five projects conducted HCW surveys. In total, 33 HCWs using 99DOTS in 4 countries and 20 HCWs using evriMED in Ukraine completed surveys (*Table 2*). Sample sizes per project ranged from 12 to 23 HCWs. Responses to open-ended survey questions were provided by 33 HCWs in Tanzania and the Philippines using 99DOTS, and 20 HCWs in Ukraine using evriMED *(Supplemental Table S5*).

**Table 2:**
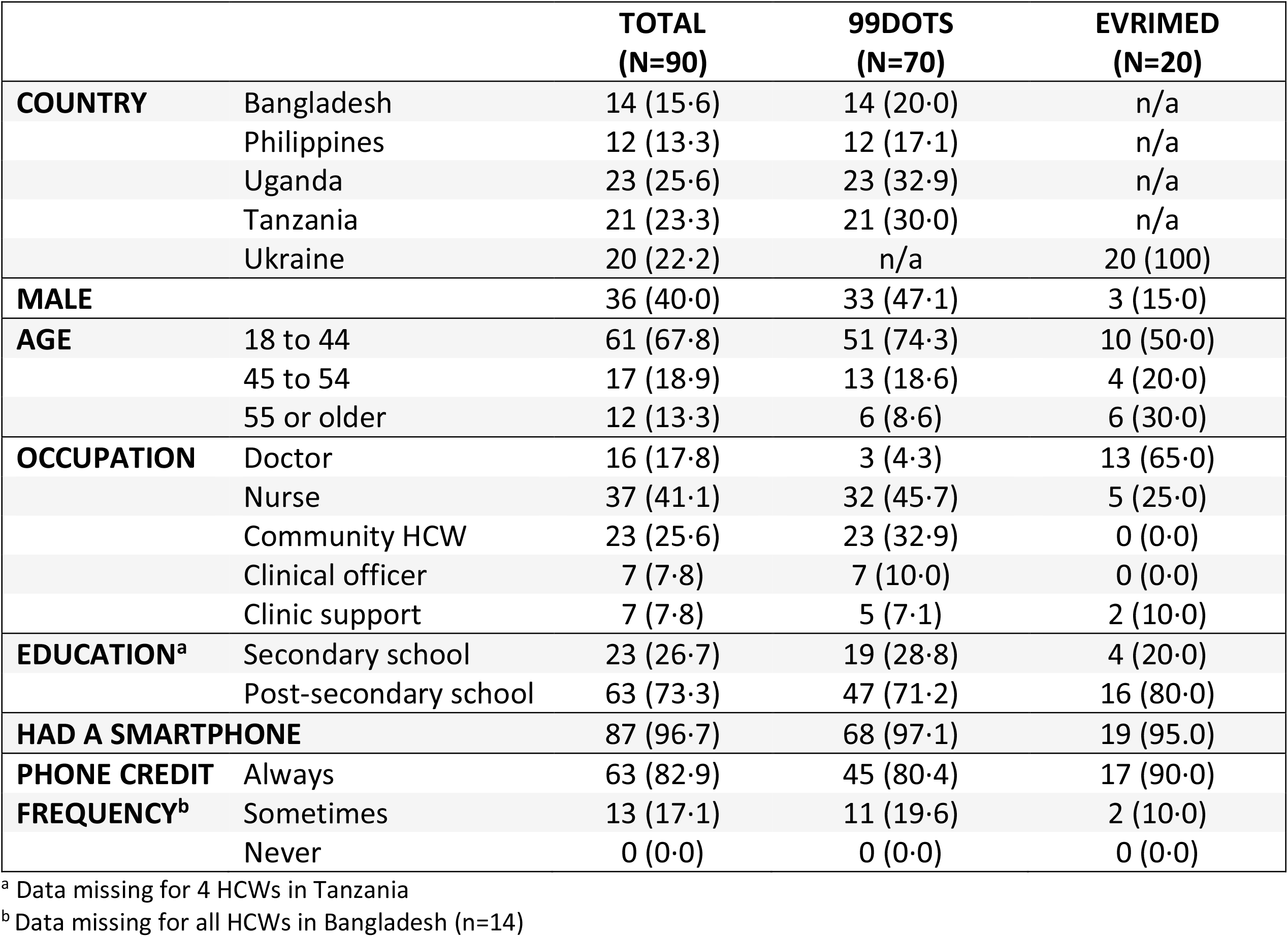
Demographic characteristics of health care workers.

### Demographics and access to technology

Across all projects, 61·3% (n=791) of people with TB were male, and 68·5% (n=884) were younger than 45 years old *(Table 1)*. Most were self-employed (n=344, 30·2%) or hired for work (n=375, 32·9%). A higher proportion of people using evriMED were unemployed compared to 99DOTS (47·5% v. 8·0%), likely reflecting differences in the settings where projects were conducted. Most had completed secondary school education (n=731, 56·5%) with a higher proportion of evriMED users completing secondary school (91·6% v. 47·4%). Almost all people with TB (n=1,072, 83·8%) had access to their own phone or were the primary owner of a shared phone. Most only sometimes had access to phone credit (n=834, 65·3%), and 13·5% (n=172) reported never having access to phone credit.

Most HCWs surveyed were nurses (n=37, 41·1%) or community health workers (n=23, 25·6%), reflecting the primary staff involved in follow-up of people receiving TB treatment (*Table 2)*. Of 90 HCWs surveyed, 54 (60%) were female and 61 (67·8%) were younger than 45 years old. All HCWs had completed at least secondary school with the majority (n=63, 73·3%) completing post-secondary school. Almost all (n=87, 96·7%) had access to a smartphone and always had phone credit (n=63, 82·9%). Some (n=13, 17·3%) reported only sometimes having phone credit and no one reported not having phone credit (*Supplemental Table S7*).

### Experience using DATs

Overall, 44·1% (n=233) of people with TB reported having taken their medication but not reporting it using DAT at least once during treatment (*Table 3)*. This was similar among those using 99DOTS and evriMED (42·8% v. 46·0%). The most common reason for not reporting taken doses were problems with charging the battery of their device, poor network connection, forgetting to use the DAT, and the DAT not working. Only 2 (0·9%) responded that they did not want to use the DAT. About one-fifth (n=132, 22·9%) of people with TB reported missing a dose of medication. Common reasons included forgetting to take their medication, medication side effect, and being too busy. They usually took less than 2 minutes to take their medication and made 1 to 5 visits a month to a TB clinic. Most (n=335, 78·3%) lived within an hour of their clinic, however 93 (21·7%) had to travel more than an hour.

**Table 3:**
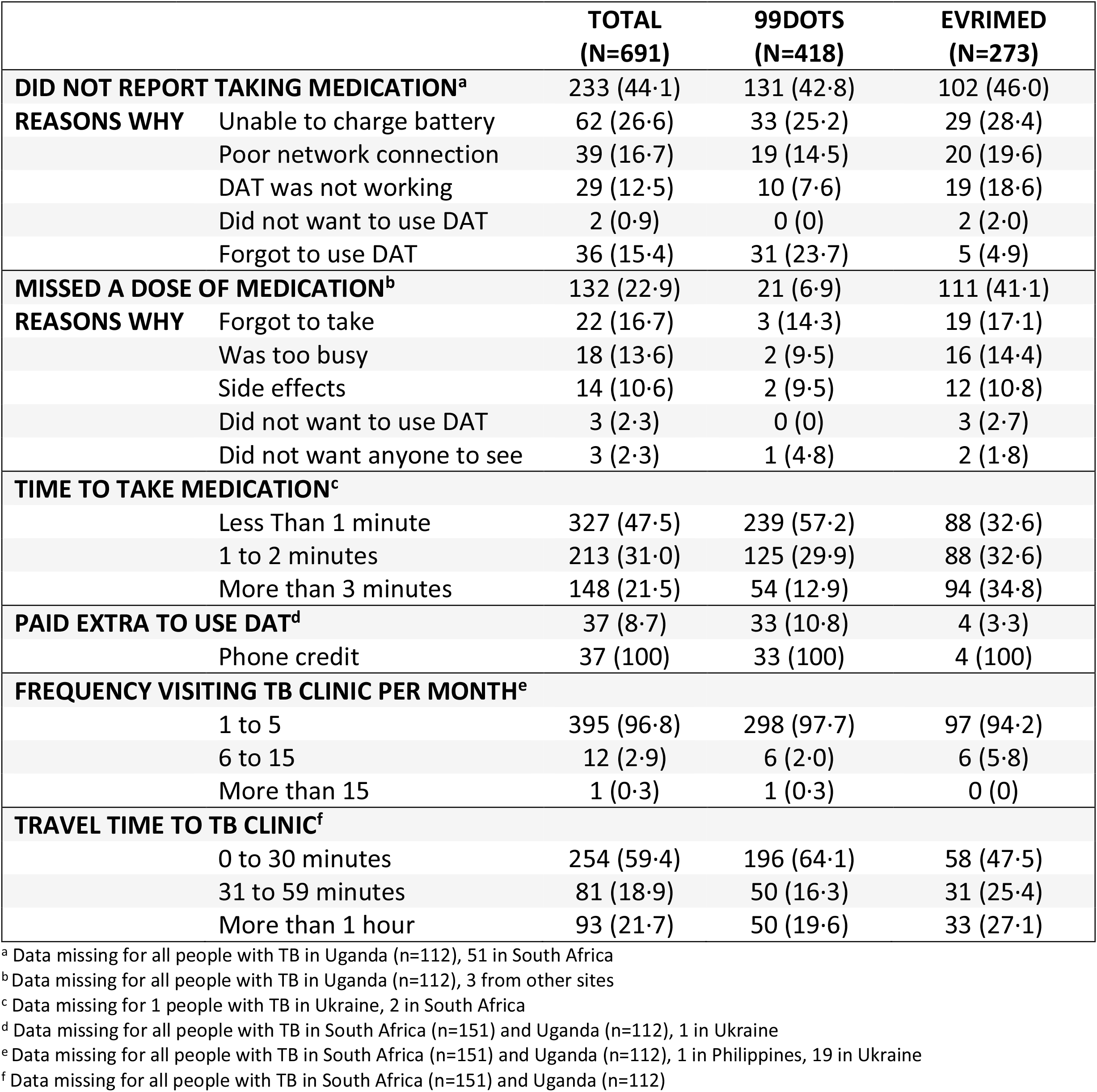
Experience using DAT among people on TB treatment.

**Table 4:**
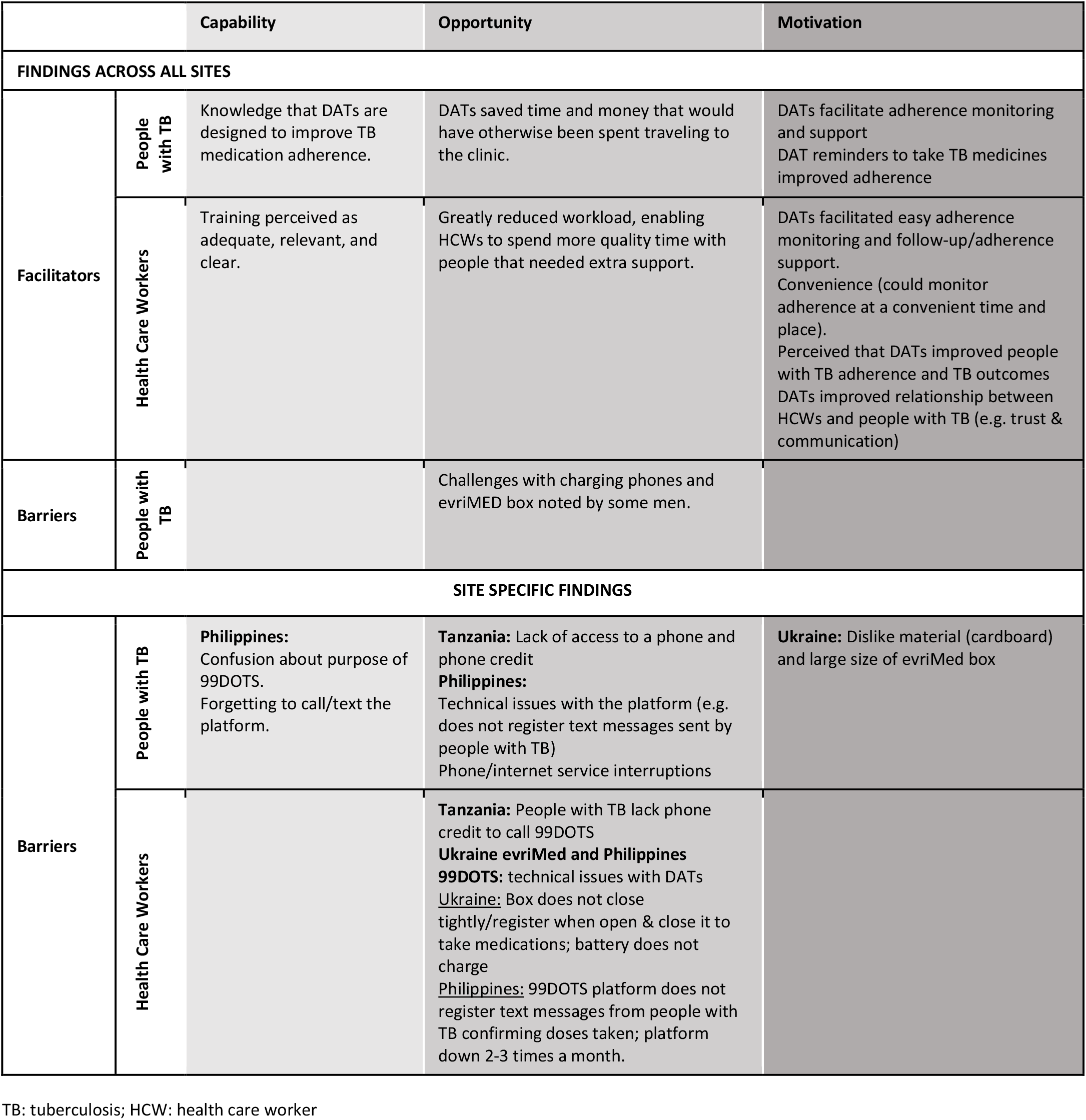
Summary of qualitative analysis of responses from people with TB and health care workers by COM-B Category.

All HCWs received data from the DAT and most reviewed data daily (n=73, 90·1%) through phone messages, mobile phone applications, and/or computers (*Supplemental Table S8)*. Adherence was most frequently assessed using the mobile phone application (n=77, 85·6%). In addition, HCWs reported calling people with TB (n=56, 62·2%), asking when they refilled medications (n=51, 56·7%), and talking to their family members (n=44, 48·9%). Throughout the project, 52·2% (n=47) of HCWs were not able to assess adherence data at some point. HCWs cited problems with poor network connection (n=38, 80·9) and the mobile phone application not working (n=34, 72·3%). Some also experienced electricity outages or limited access to records. No HCW responded that they did not want to use the DAT.

### Capability to use DAT

Overall, people with TB had a favorable impression of their capability to use DAT (*Figure 2*). Mean responses for both 99DOTS (4·49, 95% CI 3·95-5·03) and evriMED (4·58, 95% CI 3·86-5·31) were near 4·5, reflecting a response between agree and strongly agree. There were no differences in mean capability scores by DAT type, sex or age (*Supplemental Figures S1 – S2*).

**Figure 2:**
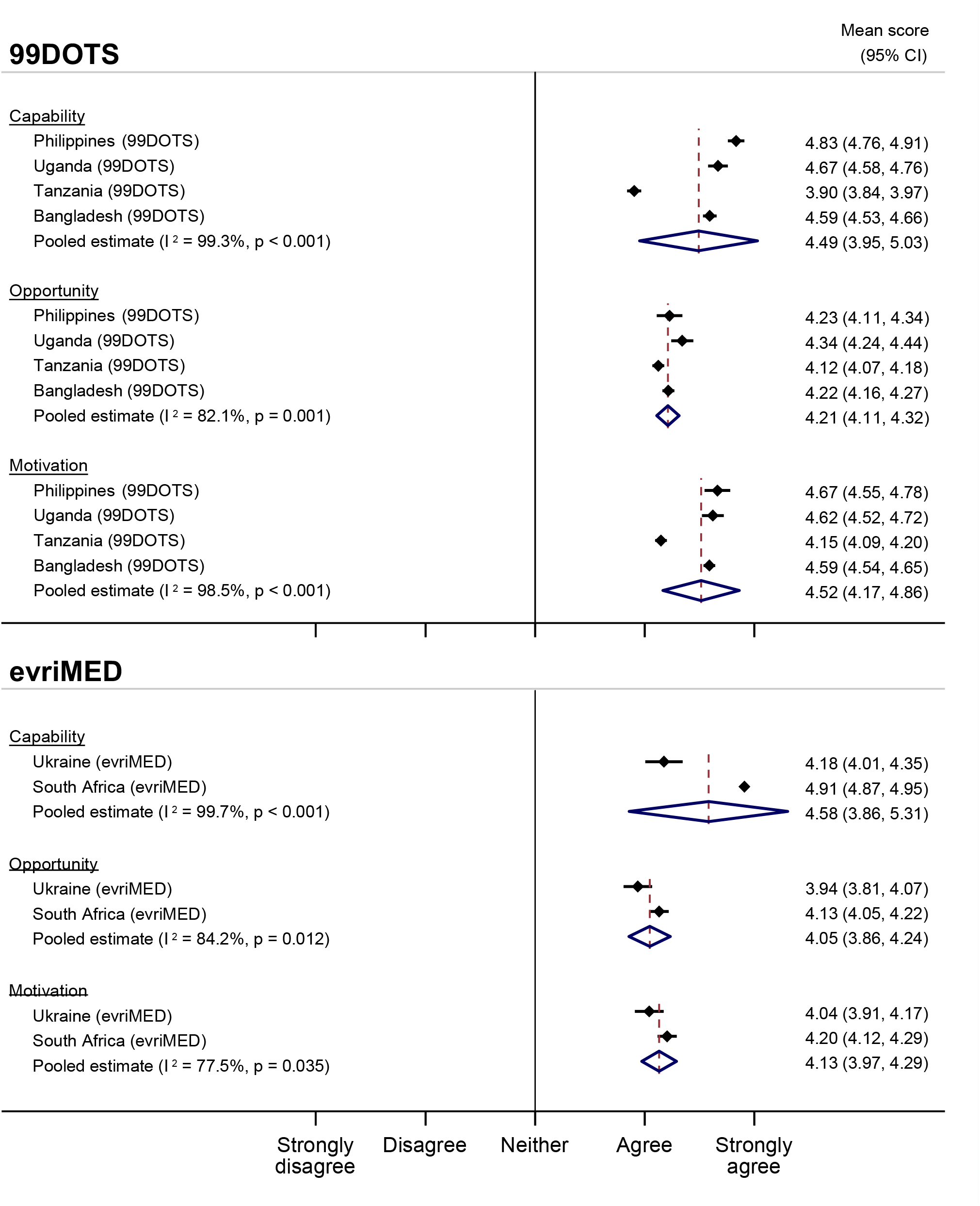
Mean Capability, Opportunity, and Motivation scores by DAT and country among people with TB.

Capability was facilitated by knowledge that the DAT was a tool to help people with TB remember to take their medication and improve their adherence. They reported that DATs reminded them to take their medications and facilitated adherence monitoring and support.

> *“The fact that with the smartbox it’s impossible to forget to take medicines. It help me to become healthier*.*”*-Female 45-54, Ukraine evriMED

Some in the Philippines (mostly men) described forgetting to call/text the platform and confusion about the purpose of 99DOTS:

> *“With 99DOTS, sometimes I just really forget to text. I’d sometimes tell myself I’d do it later, but I keep forgetting”*-Male, 15-44, Philippines 99DOTS

Overall, HCWs also had a favorable impression of their capability to use DAT *(Figure 3)*. HCWs in Tanzania had exclusively strongly agree responses in the capability domain. Pooled estimates for HCWs using 99DOTS (4·89, 95% CI 4·79-4·98) and evriMED (4·78, 95% CI 4·65-4·90) were both high. HCWs felt that the training they received was relevant, useful, and clear, facilitating their capability to successfully implement DATs. HCWs suggested refresher training on any updates to the device/platform would further improve their capability.

**Figure 3:**
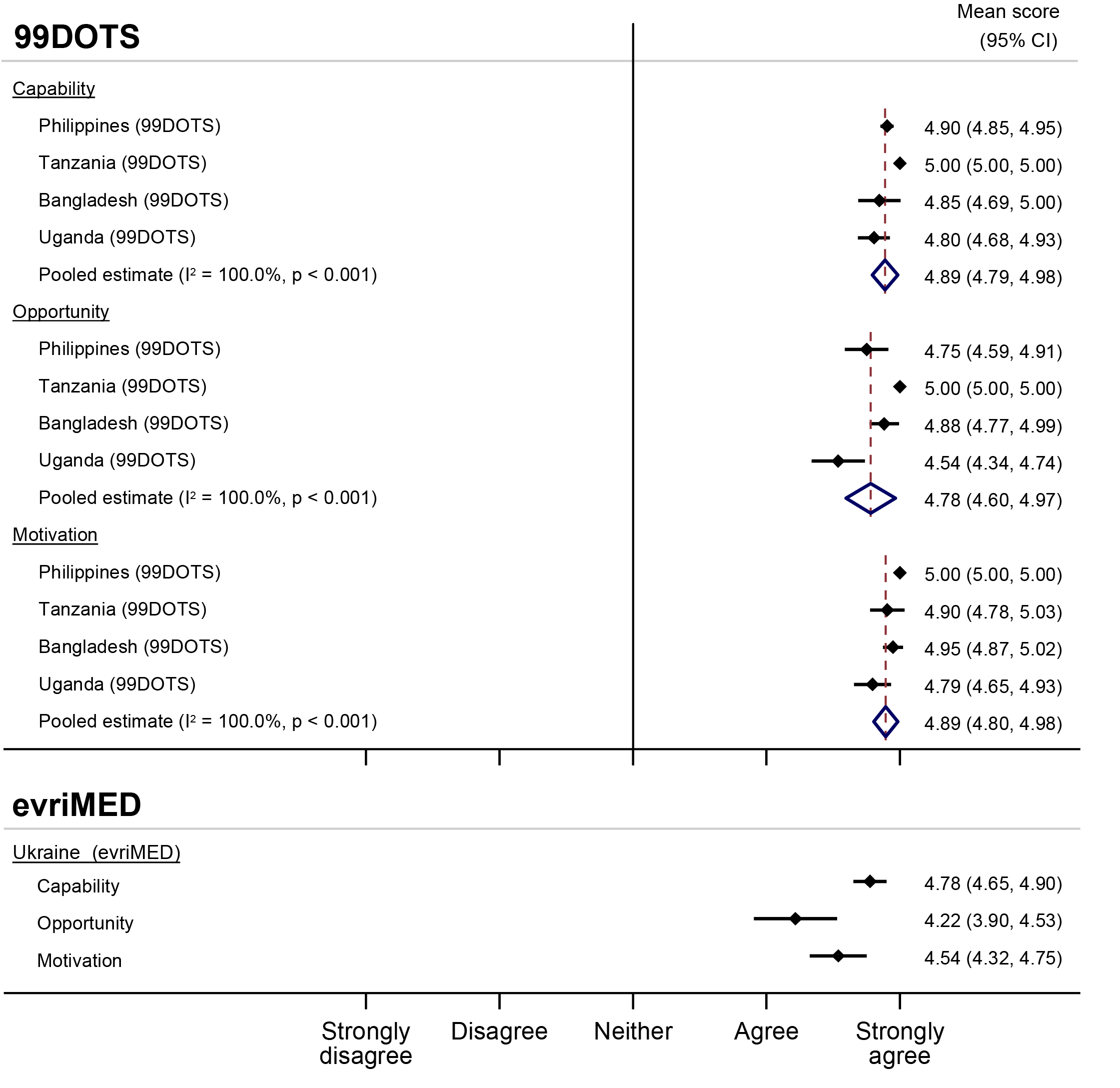
Mean Capability, Opportunity, and Motivation scores by DAT and country among health care workers.

### Opportunity

Within opportunity, people with TB largely thought that using DAT was acceptable (did not take too much time, increased connection with HCWs) and that they would recommend it to their family members. But there were some concerns related to using DAT in front of others as it could result in the disclosure of TB status (Figure 2). Pooled estimates for 99DOTS (4·21, 95% CI 4·11 – 4·32) and evriMED (4·05, 95% CI 3·86 – 4·24) were less favorable than for capability, but overall still positive with responses averaging above “agree”. There were no differences by gender or age (*Supplemental Figures S3 – S4)*.

People with TB across all studies reported that DATs saved time and money that they would have otherwise spent traveling to clinic each day to take their TB medication under DOT:

> *“With the smartbox I can stay at home. It saves my time and money*.*”-*Male, 65-74, Ukraine evriMED

Those using 99DOTS also liked that calling or texting 99DOTS was free and noted greater social support:

> *“I get treated for TB for free and I have to text 99DOTS everyday. My eldest reminds me too and asks me if I’ve already taken my medicine for the day*.*”*-Female, 45-54, Philippines 99DOTS

People using both 99DOTS and evriMED reported issues with charging their phones or boxes. Those using evriMED disliked the design of the box, pointing to the cardboard material and large size. In Tanzania, where calling the 99DOTS platform was not toll-free, nearly half of participants indicated lack of access to a phone and phone credit as major barriers to using 99DOTS.

Responses by HCW also showed favorable opportunity to use DATs (*Figure 3*). HCWs using 99DOTS in Tanzania exclusively had “strongly agree” responses to questions in the opportunity domain. Pooled estimates were high for both 99DOTS (4·22, 95% CI 3·90 – 4·53) and evriMED (4·78, 95% CI 4·60 – 4·97).

HCWs endorsed decreased workloads and thought that DATs were more person-centered, relieving some of the financial impact of TB treatment and allowing people with TB to go on long trips and continue working. Several HCWs noted that DATs enabled them to spend more quality time engaging with people with TB who needed their support or to attend to other administrative tasks. Barriers included difficulty calling people with TB if they did not have phone credit and lack of access to phones in Tanzania. Additionally, there were technical issues with the evriMED box not closing tightly or not registering when they took their medication and 99DOTS not registering all text messages from them. A small group of HCWs (primarily from Ukraine) perceived DATs increased their workload by adding more forms and procedures for them to complete each day.

### Motivation

People with TB were motivated to complete treatment and get healthy, but there were concerns about data privacy. Motivation was favorable for using both 99DOTS (pooled estimate 4·52, 95% CI 4·17-4·86) and evriMED (pooled estimate 4·52, 95% CI 4·17-4·86) (*Figure 2*). There were no differences in motivation by gender or age *(Supplemental Figures S5 – S6)*.

HCWs also demonstrated high motivation to use DAT, having almost exclusively “strongly agree” responses for both 99DOTS (pooled estimate 4·89, 95% CI 4·80-4·98) and evriMED (pooled estimate 4·54, 95% CI 4·32-4·75) (*Figure 3)*. Across projects, HCWs described liking that DATs facilitated easy adherence monitoring and follow-up/adherence support, and believed that DATs improved adherence for people with TB.

## Discussion

Overall, we found that 99DOTS and evriMED were both highly acceptable to people receiving treatment for DS-TB and HCWs caring for them. However, there were barriers encountered that decreased the feasibility of DATs, some of which can be targets for future optimization of these technologies and their implementation. Neither age nor gender had an impact on the perceived acceptability or feasibility of the two DAT platforms.

Our findings largely support prior studies demonstrating DATs offer an acceptable, person-centered approach to TB care.^21–23^ High acceptability was demonstrated by high capability, opportunity, and motivation to use 99DOTS and evriMED among people with TB and HCWs. Both agreed that DATs saved time and money which would have otherwise been spent traveling to clinics. Our study also found that people with TB using DAT felt strongly connected to their HCWs. Most HCWs endorsed reduced workload when using DAT. Thomas et al reported similar findings among HCWs at government-run TB centers in India.^24^ Stigma related to taking TB medication contributed to lower opportunity scores, reflecting the known impact of stigma on TB care.^25^

In addition to stigma, barriers to the feasibility of implementing DATs included technical issues with phones, network connectivity, and issues with the two DAT platforms. Although access to cell phones among people with TB was high, 44% reported having taken their TB medication but not reporting it using their DAT. Issues with charging phones/boxes and network connectivity were barriers to using DATs across all projects. In Tanzania, the only 99DOTS project where a toll-free number still required phone credit, nearly half of people with TB completing open-ended questions mentioned the cost associated with needing phone credit as a key barrier. A minority reported other issues with their DAT, such as the evriMED box not closing tightly and the 99DOTS platform not registering texts or calls that were sent from people with TB. Similar technical barriers have been reported previously in China, India, Peru, and sub-Saharan Africa.^24,26–29^

Our analysis had some limitations. First, surveys were administered only to a subset of people with TB in each project. Selection bias is therefore possible and could have led to more favorable responses. Second, since the surveys were implemented by the project teams, participating people with TB and HCWs may have felt pressure to please the person administering the survey. Social desirability bias may result in more favorable responses. Survey administration skills also varied between projects. Third, selection of DAT was not randomized, which prevented direct comparison of acceptability and feasibility of the two DAT platforms; differences observed may be due to setting or project implementation. Finally, qualitative data was only collected via open-ended survey questions and not through in-depth interviews. This may have prevented us from gaining a deeper and more nuanced understanding of the barriers and facilitators to DAT implementation and acceptability.

In summary, the available data show high acceptability of DAT and support further scale-up of DAT for treatment of DS-TB. However, implementation efforts should focus on ensuring access, anticipating and addressing technical challenges with the platforms, and minimizing additional costs to people with TB. Further research comparing DAT platforms in the same settings is needed to better inform whether certain platforms are more acceptable and feasible to use for specific populations.

## Data Availability

All data produced in the present study are available upon reasonable request to the authors
All data produced in the present work are contained in the manuscript

## Contributors

The study was conceived by AK and AC. Data collection was conducted by NM, LJ, BO, LS, JA, JL, SA, SS, AK, AK, KG, and AB. Data cleaning, analysis, interpretation, and validation was done by KG, RC, AL, with oversight from AC and AK. KG, RC, and AL wrote the first draft of the manuscript and AK and AC reviewed and revised the manuscript. All authors contributed to and approved the final manuscript.

## Data Sharing

The anonymized datasets used in this study can be available upon reasonable request to the corresponding authors.

## Acknowledgments

We are grateful to all the people with TB and the health care workers who participated in the survey. We also would like to acknowledge Miranda Brouwer, Danielle Cazabon, Kristian Van Kalmthout, Tripti Pande, and Oriol Ramis for helping conceptualize this work or having input into the data collection tools.

## Supplemental Material

**Table S1:** Constructs assessed in survey administered to people with TB by COM-B category.

| TDF construct | Statement |
| --- | --- |
| **Capability** |  |
| Knowledge | I know how to use DAT |
| Memory, attention, and decision processes | The reminders I receive on my phone help me to remember to take my TB medicines |
| **Opportunity** |  |
| Social influences | I am worried that using DAT makes it more likely others will find out I have TB |
|  | I am comfortable using DAT in front of other people |
|  | I am comfortable using DAT outside of my home |
|  | I will recommend using DAT to my family or friends if they have TB |
|  | DAT helps me feel more connected to my health workers |
| Environmental context and resources | It takes me too much time to take my TB medicines and call DAT every day |
|  | I make fewer trips to the health clinic for my TB treatment because I am using DAT |
| **Motivation** |  |
| Optimism | Using DAT will help me to complete my TB treatment |
|  | Using DAT while taking TB medicines will help me get healthy |
| Reinforcement | The DAT packaging makes it easy for me to remember what to do |
| Emotion | I am concerned about the privacy of my health information collected by the DAT system |

COM-B: Capability Opportunity Motivation Behavior; TDF: Theoretical Domains Framework; DAT: digital adherence technology; TB: tuberculosis

**Table S2:** Constructs assessed in survey administered to health care workers, by COM-B category.

| **TDF construct** | **Statement** |
| --- | --- |
| **Capability** |  |
| Knowledge | I understand the DAT system and how it works<br>I received adequate training to use DAT |
| Memory, attention and decision processes | I remember to check the adherence data available in DAT when patients come in for refill visits<br>The reminders I receive from DAT help me to remember to check on patients who are not taking their medicines |
| Effort Expectancy* | It is easy for me to explain how to use DAT to my patients<br>It is easy for me to identify which patients are not taking their TB medicine using DAT |
| **Opportunity** |  |
| Social influence^ | My patients like using DAT |
| Effort Expectancy* | Learning to use DAT is easy for me<br>It is easy for me or my co-workers to contact patients who have not taken their TB medicines<br>Using DAT helps to reduce my workload |
| **Motivation** |  |
| Belief about consequences | Using DAT improves the care I provide to my patients<br>DAT adherence data helps me provide better support and counseling to my patients |
| Trust factor* | I believe that DAT data accurately reflects if my patients took their TB medicines or not |
| Purchase intention* | I would recommend using DAT to my patients |

\*Key construct of UTAUT
^Both a key construct of UTAUT and a TDF domain
COM-B: Capability Opportunity Motivation Behavior; TDF: Theoretical Domains Framework;
UTAUT: Unified Theory of Acceptance and Use of Technology; DAT: digital adherence technology; TB: tuberculosis

**Table S3:** Characteristics of studies included in the meta-analysis.

| Country | DAT | TB | Study Period | Data Collection | Population | Project Summary |
| --- | --- | --- | --- | --- | --- | --- |
| Bangladesh | 99DOTS | DS-TB | 2019-2020 | 09/2020-10/2020 | Private Clinics | Digital monitoring of TB treatment adherence for differentiated care in private provider settings. |
| South Africa | evriMED | DS-TB | 2019-2020 | 03/2020-05/2020 | Public Clinics | Randomized control trial comparing passive evriMED box v. real time monitoring of evriMED and differentiated care based on dosing histories and level of missed doses. |
| Philippines | 99DOTS | DS-TB | 2019-2020 | 11/2019-02/2020 | Private Clinics | 99DOTS as a platform for quality TB treatment by private providers where 50% of patients in the country seek care. |
| Tanzania | 99DOTS | DS-TB | 2019-2020 | 10/2019-03/2020 | Public Clinics | Empowering TB patients in a mining population using treatment adherence technologies |
| Ukraine | evriMED | DS-TB and DR-TB | 2019-2020 | 09/2019-10/2019 | Public Clinics | Practicality, cost, and impact of combining real-time evriMED monitoring for enhanced counseling of DS-TB and DR-TB patients. |
| Uganda | 99DOTS | DS-TB | 2019-2020 | 12/2019-02/2020 | Public Clinics | From DAT to DAT: An evaluation of 99DOTS to improve TB adherence in Uganda |

**Table S4:** TB REACH Wave 6 digital adherence technology projects included.

| Project | DAT | Type of population | Quantitative survey results |  | Qualitative (open-ended) survey results |  |
| --- | --- | --- | --- | --- | --- | --- |
|  |  |  | People with TB | Health care workers | People with TB | Health care workers |
| Ukraine (PATH) | evriMED | Both | X | X | X | X |
| Philippines (KNCV) | 99DOTS | Susceptible | X | X | X | X |
| Tanzania (KNCV) | 99DOTS | Susceptible | X | X | X | X |
| Uganda (UCSF) | 99DOTS | Susceptible | X | X |  |  |
| Bangladesh (icddr,b) | 99DOTS | Susceptible | X | X |  |  |
| South Africa (Aurum) | evriMED | Susceptible | X |  |  |  |

**Table S5:** People with TB and health care workers included in qualitative analyses.

| Country | Ukraine | Tanzania | Philippines |
| --- | --- | --- | --- |
| DAT evaluated | evriMED | 99DOTS | 99DOTS |
| Number of people with TB | 19 | 200 | 106 |
| Number of HCW | 20 | 21 | 12 |

**Table S6.** Demographic characteristics of people with TB included in qualitative analysis.

|  | TOTAL<br>(N=325) | 99DOTS<br>(N=306) | EVRIMED<br>(N=19) |
| --- | --- | --- | --- |
| **COUNTRY** |  |  |  |
| Ukraine | 19 (5.8) | n/a | 19 (100) |
| Tanzania | 200 (61.5) | 200 (65.4) | n/a |
| Philippines | 106 (32.6) | 106 (35.6) | n/a |
| **MALE** | 225 (69.2) | 215 (70.3) | 10 (52.6) |
| **AGE** |  |  |  |
| 15 to 44 | 199 (61.2) | 188 (61.4) | 11 (57.9) |
| 45 to 64 | 101 (31.1) | 94 (30.7) | 7 (36.8) |
| 65 or older | 25 (7.7) | 24 (7.8) | 1 (5.3) |

**Table S7.** Demographic characteristics of health care workers included in qualitative analysis.

|  | **TOTAL**<br>**(N=53)** | **99DOTS**<br>**(N=33)** | **EVRIMED**<br>**(N=20)** |
| --- | --- | --- | --- |
| **COUNTRY** |  |  |  |
| Ukraine | 20 (38) | n/a | 20 (100) |
| Tanzania | 21 (39) | 21 (64) | n/a |
| Philippines | 12 (22) | 12 (36) | n/a |
| **FEMALE** | 34 (63) | 17 (52) | 17 (85) |
| **AGE** |  |  |  |
| 18 to 44 | 31 (58.5) | 20 (60.6) | 11 (55.0) |
| 45 to 64 | 20 (37.7) | 13 (39.4) | 7 (35.0) |
| 65 or older | 2 (3.8) | 0 (0) | 2 (10.0) |
| **OCCUPATION** |  |  |  |
| Doctor | 19 (35) | 6 (18) | 13 (62) |
| Nurse | 21 (39) | 16 (48) | 5 (25) |
| Counselor | 2 (4) | 2 (6) | 0 (0) |
| Pharmacist | 1 (2) | 1 (3) | 0 (0) |
| Other | 10 (19) | 8 (24) | 2 (10) |

**Table S8.** Experience using digital adherence technologies among health care workers.

|  | **TOTAL**<br>**(N=90)** | **99DOTS**<br>**(N=70)** | **EVRIMED**<br>**(N=20)** |
| --- | --- | --- | --- |
| **PEOPLE WITH TB SEEN PER DAY** |  |  |  |
| 1 to 2 | 8 (8·9) | 7 (10·0) | 1 (5·0) |
| 3 to 5 | 34 (37·8) | 28 (40·0) | 6 (30·0) |
| 6 to 10 | 18 (20·0) | 16 (22·9) | 2 (10·0) |
| 10 or more | 30 (33·3) | 19 (27·1) | 11 (55·0) |
| **RECEIVES DATA FROM DAT** | 90 (100) | 70 (100) | 20 (100) |
| **DATA FREQUENCY <sup>A</sup>** |  |  |  |
| Daily | 73 (90·1) | 56 (91·8) | 17 (85·0) |
| 3 to 5 times per week | 7 (8·6) | 5 (8·2) | 2 (10·0) |
| Less than 3 times per week | 1 (1·2) | 0 (0·0) | 1 (5·0) |
| **DATA TYPE** |  |  |  |
| Phone message | 50 (55·6) | 39 (55·7) | 11 (55·0) |
| Mobile app | 39 (43·3) | 23 (32·9) | 16 (80·0) |
| Computer | 12 (13·3) | 7 (10·0) | 5 (25·0) |
| Tablet | 18 (20·0) | 18 (25·7) | 4 (20·0) |
| Call from a technician | 9 (10·0) | 9 (12·9) | 0 (0·0) |
| **WAYS THAT ADHERENCE WAS ASSESSED** |  |  |  |
| Ask when they refill meds | 51 (56·7) | 41 (58·6) | 10 (50·0) |
| Count pills | 21 (23·3) | 21 (30·0) | 0 (0·0) |
| Ask them to come to clinic | 12 (13·3) | 4 (5·7) | 8 (40·0) |
| Talk to family | 44 (48·9) | 32 (45·7) | 12 (60·0) |
| Talk on the phone | 56 (62·2) | 37 (52·9) | 19 (95·0) |
| Use mobile app | 77 (85·6) | 63 (90·0) | 14 (70·0) |
| **NOT ABLE TO ACCESS DAT TO ASSESS ADHERENCE** | 47 (52·2) | 41 (58·6) | 6 (30·0) |
| **REASONS WHY** |  |  |  |
| No electricity | 5 (10·6) | 4 (9·8) | 1 (16·7) |
| Poor network connection | 38 (80·9) | 34 (82·9) | 4 (66·7) |
| App was not working | 34 (72·3) | 33 (80·5) | 1 (16·7) |
| Did not want to use DAT | 0 (0·0) | 0 (0·0) | 0 (0·0) |
| Limited access to records | 8 (17·0) | 8 (19·5) | 0 (0·0) |

<sup>a</sup> Data missing for 8 HCWs in the Philippines, 1 HCW in Tanzania

**Table S9.** Internal consistency of survey responses among people with TB.

|  | Capability* | Opportunity* | Motivation* |
| --- | --- | --- | --- |
| **99DOTS** | 0.45 | 0.58 | 0.54 |
| **evriMED** | 0.21 | 0.53 | 0.55 |
| **Overall** | 0.38 | 0.36 | 0.56 |

\* Cronbach's alpha is shown in the data cells

**Table S10.**
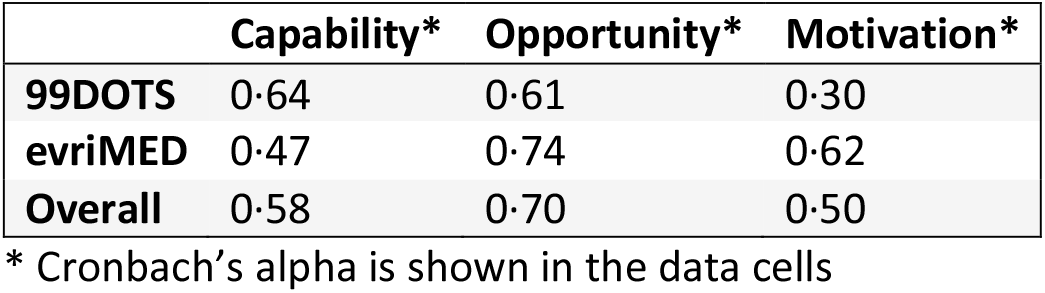
Internal consistency of survey responses among health care workers.

|  | Capability* | Opportunity* | Motivation* |
| --- | --- | --- | --- |
| **99DOTS** | 0.64 | 0.61 | 0.30 |
| **evriMED** | 0.47 | 0.74 | 0.62 |
| **Overall** | 0.58 | 0.70 | 0.50 |

\* Cronbach's alpha is shown in the data cells

**Figure S1.**
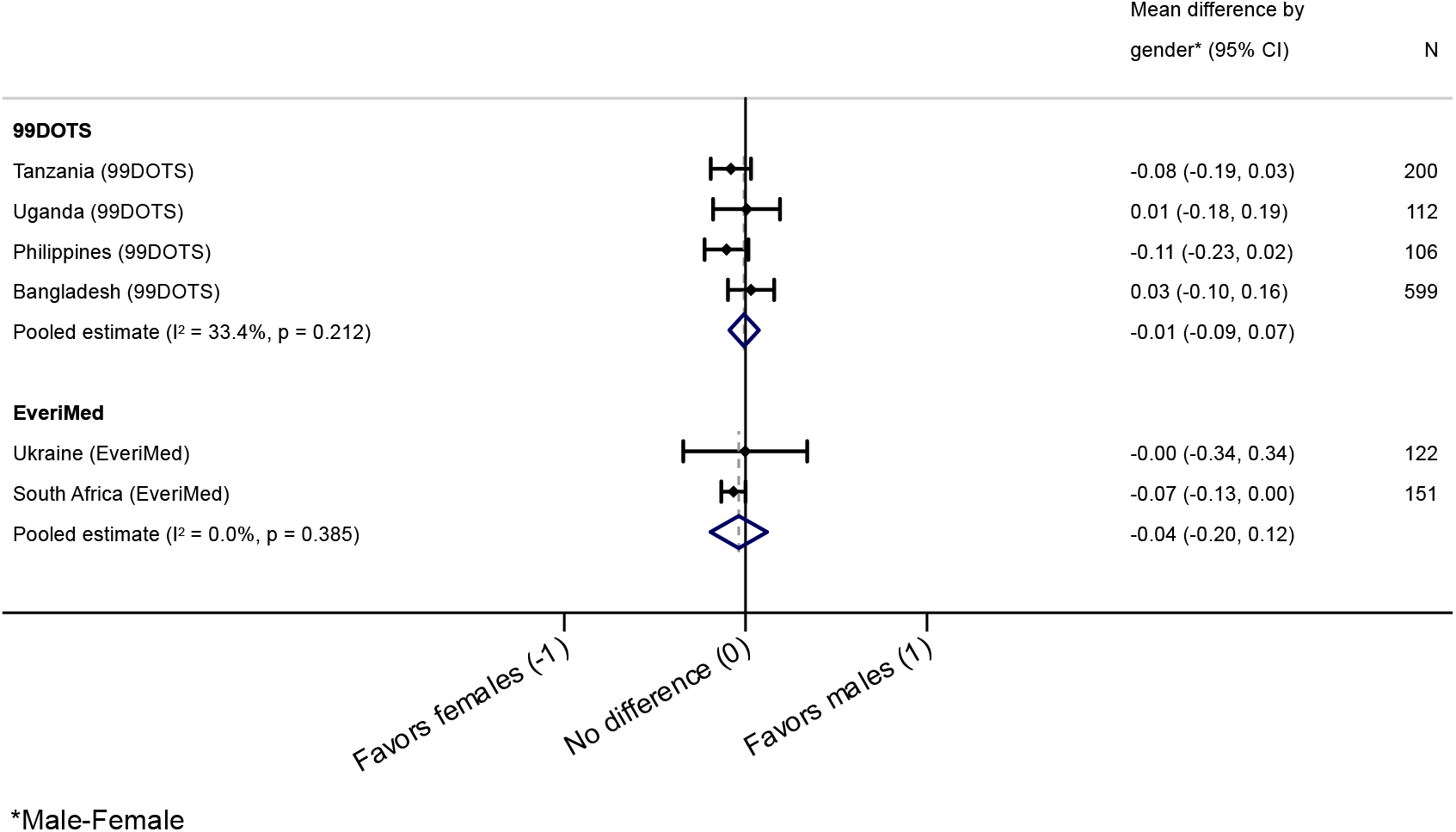
Difference in mean Capability scores by gender among people with TB.

**Figure S2.**
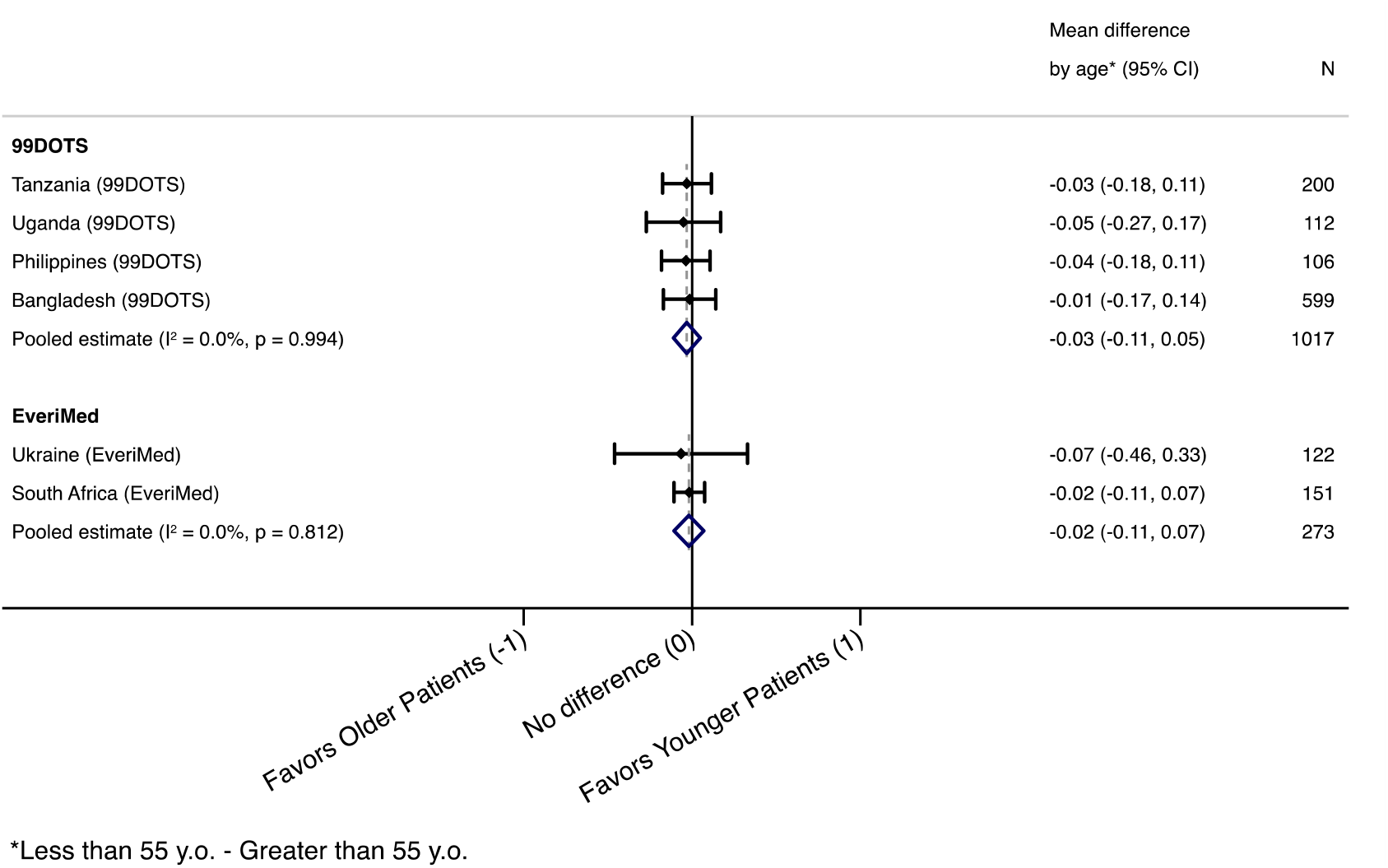
Difference in mean Capability scores by age (≥55 vs. <55 years) among people with TB.

**Figure S3.**
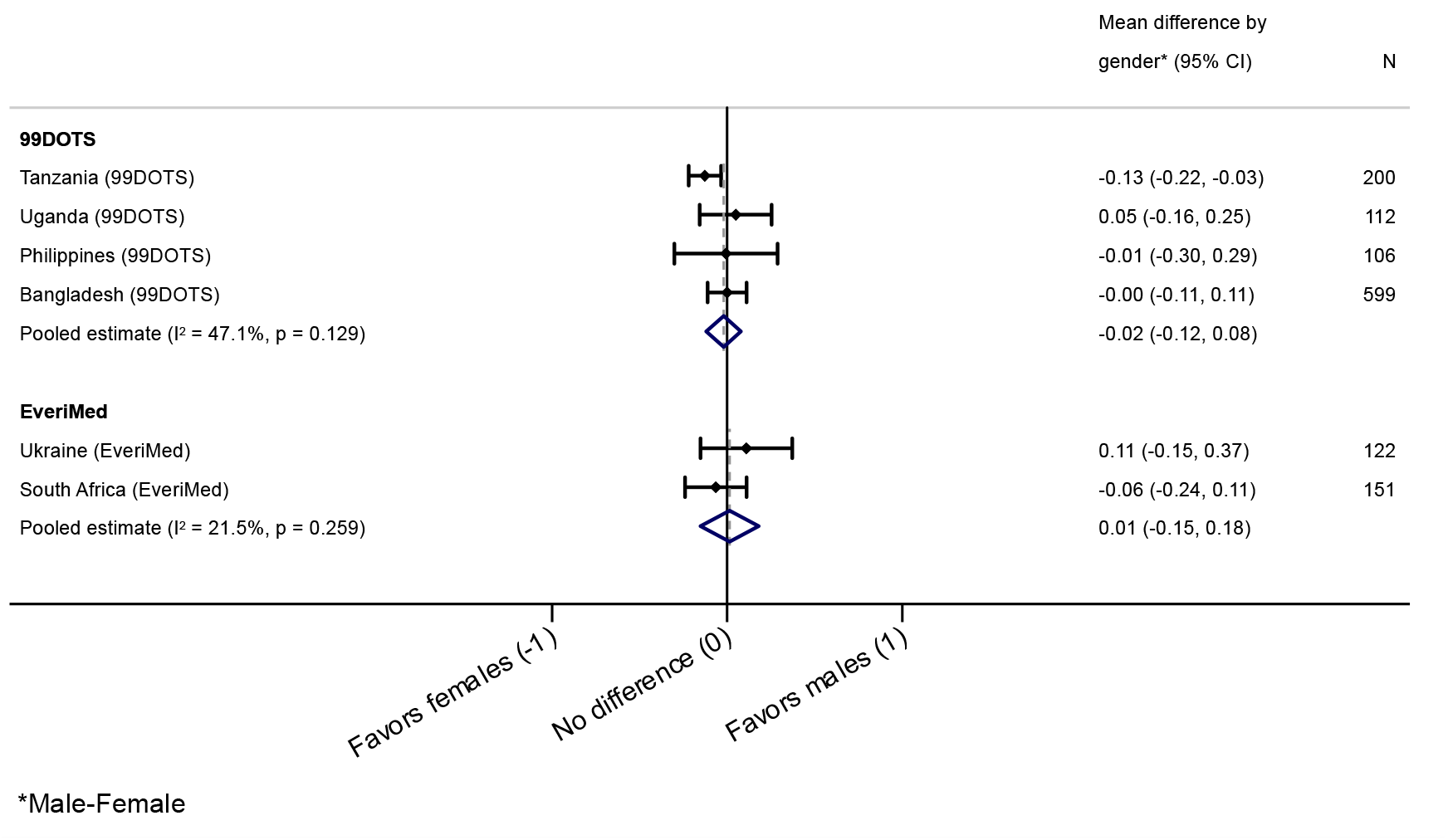
Difference in mean Opportunity scores by gender among people with TB.

**Figure S4.**
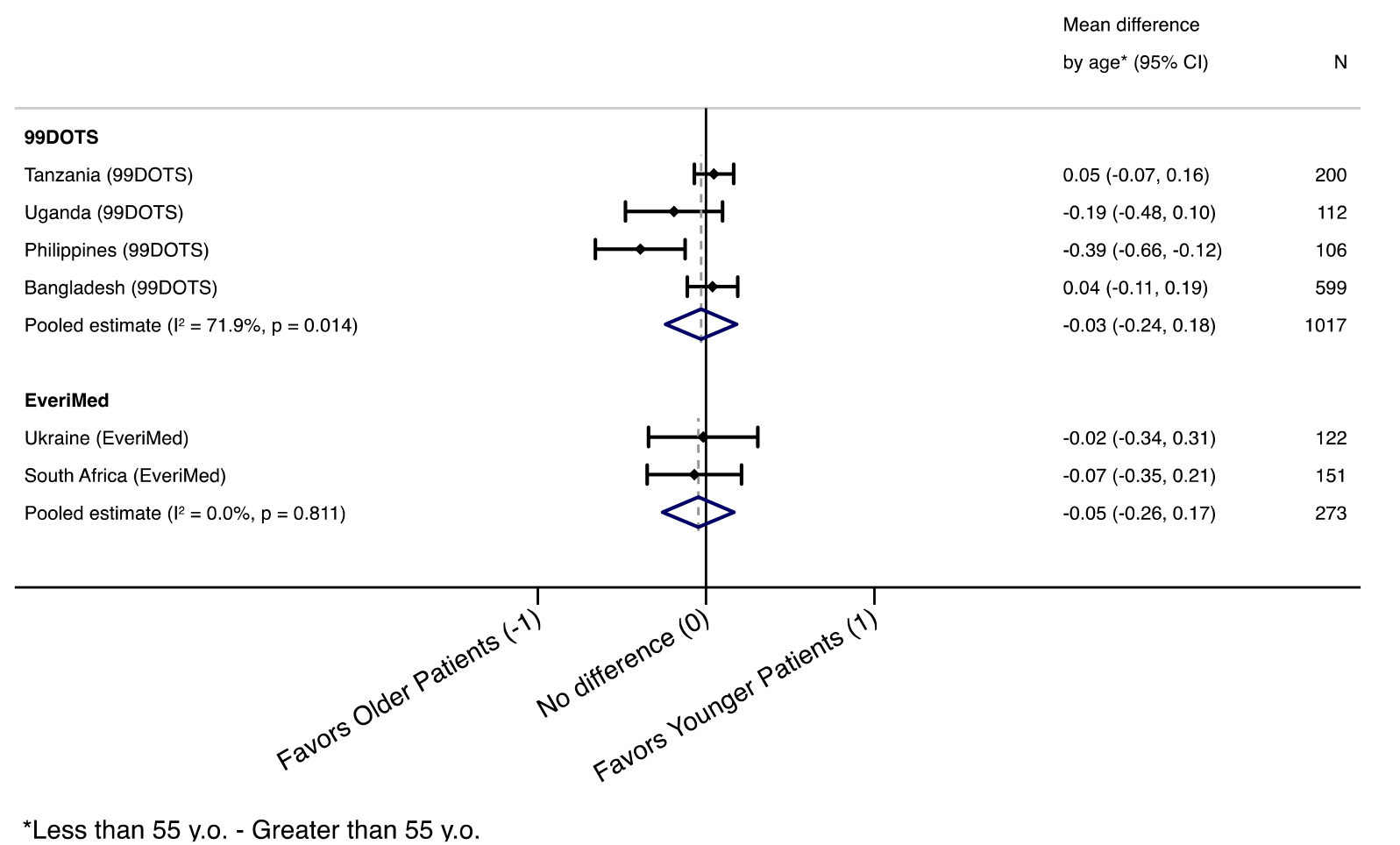
Difference in mean Opportunity scores by age (≥55 vs. <55 years) amoung people with TB.

**Figure S5.**
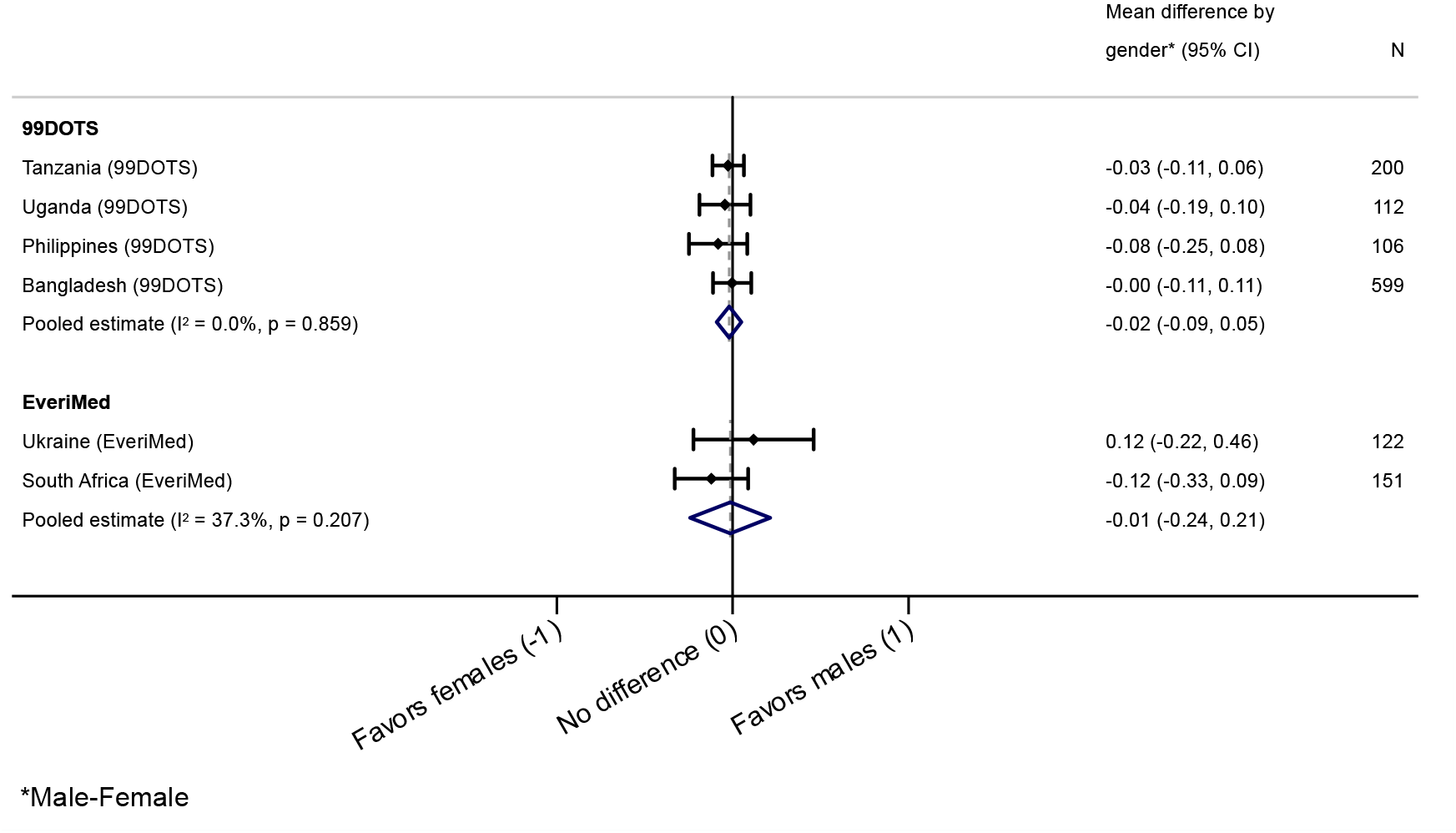
Difference in mean Motivation scores by gender among people with TB.

**Figure S6.**
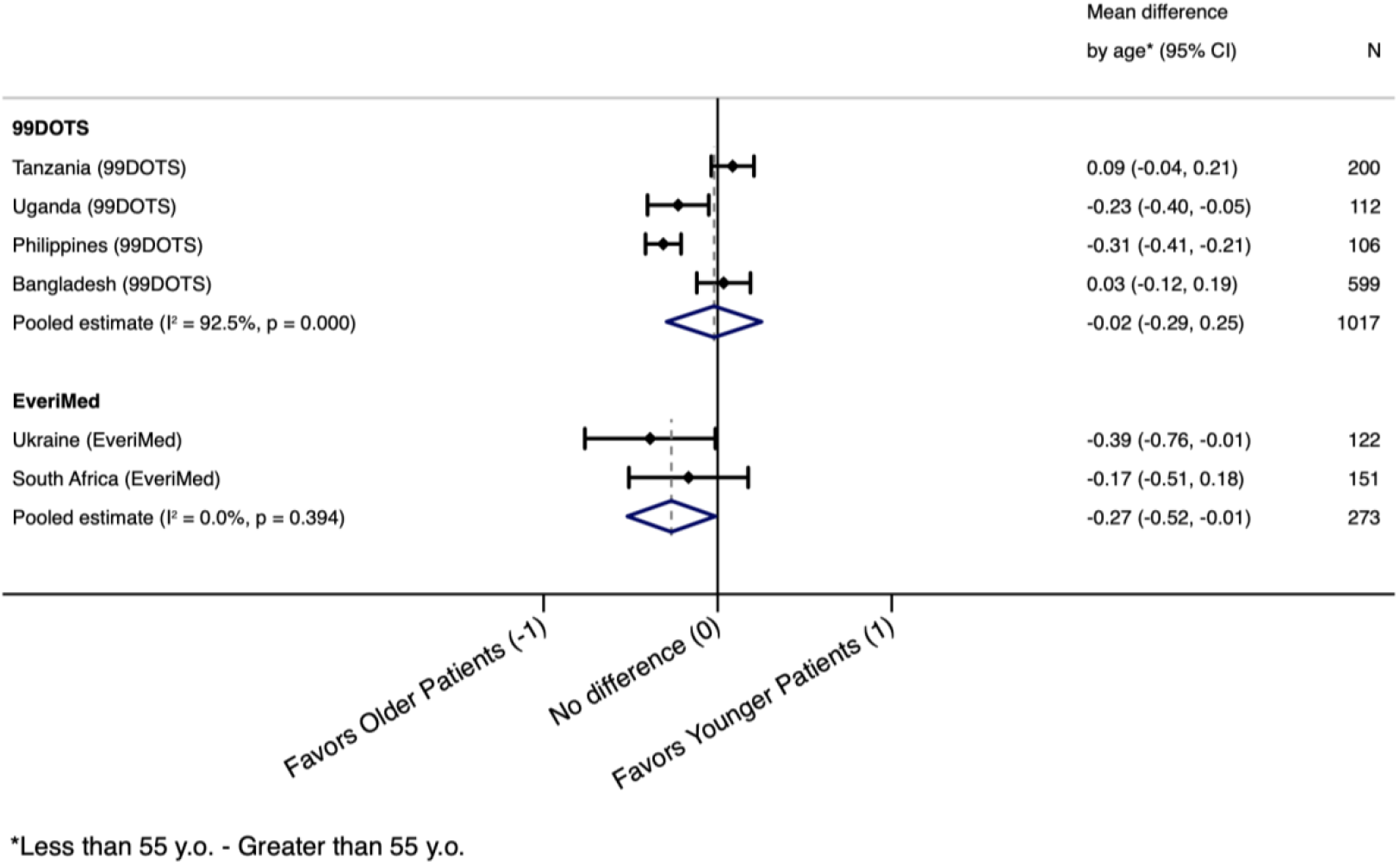
Difference in mean Motivation scores by age (≥55 vs. <55 years) among people with TB.

## Abbreviations

COM-B: Capability, Opportunity, Motivation, Behavior
BMGF: Bill and Melinda Gates Foundation
DAT: Digital adherence technology
DR-TB: Drug-resistant TB
DS-TB: Drug-sensitive TB
GAC: Global Affairs Canada
HCW: Health care workers
TB: Tuberculosis
TDF: Theoretical Domains Framework
UTAUT: Unified Theory of Acceptance and Use of Technology
VDOT: Video directly observed therapy
WHO: World Health Organisation

